# Adaptation and validation of a French version of the Vaccination Attitudes Examination (VAX) scale

**DOI:** 10.1101/2022.04.28.22274372

**Authors:** Margot Eisenblaetter, Clarisse Madiouni, Yasmine Laraki, Delphine Capdevielle, Stéphane Raffard

**Author notes:** Corresponding Author: Margot Eisenblaetter.

## Abstract

Over the past decades, vaccination has proven to be largely beneficial to global health. Despite vaccine efficacy, the French population has been recently affected by more anti-vaccination attitudes and vaccine refusal, and it is therefore necessary to develop and validate tools to study this health issue. The Vaccination Attitudes Examination scale is a brief 12-item questionnaire targeting adults, that assesses general attitudes towards vaccination. The aims of the study were (1) to translate and adapt the original English version of the scale into French and (2) to test the psychometric properties of the scale in a French population-based sample of adults. We included 450 French speaking adults that completed the French Vaccination Attitudes Examination scale and other questionnaires in order to assess convergent and divergent validities. Exploratory and confirmatory factor analyses showed that the French version of the Vaccination Attitudes Examination scale replicated the factorial structure of the original scale. Moreover, it demonstrated high internal consistency, good convergent and divergent validities, and excellent temporal stability. Also, scores on the scale differentiated vaccinators from non-vaccinator respondents. Results on the scale provides us with insight into factors involved in vaccine hesitancy in France, therefore allowing French authorities and policy makers to address these specific concerns and by consequence improve vaccine acceptance rates in this country.

## Introduction

Vaccination has been the one most effective measure to prevent and control many infectious diseases and has proven to be largely beneficial to global health ^[1]^. For instance, smallpox (in 1979) and rinderpest virus (in 2011) are two infectious diseases that have been eradicated by vaccination. The World Health Organization (WHO) established in 1974 a successful programme aiming to facilitate vaccination uptake for children all around the world. This programme allowed to radically reduce death rates due to infectious diseases beyond borders of developing countries ^[2]^. As a result, the WHO estimated that vaccination has saved 2-3 million children each year from life-threatening diseases. Vaccination has also substantial health related benefits in adult populations ^[3]^. A study led in 2019 stated that among the 42 European countries studied, all had established national vaccination programmes for adults ^[4]^. Most of these programmes included vaccinations for influenza, diphtheria, tetanus, and pneumococcus. For instance, influenza vaccination has proven to be beneficial for healthy working adults in allowing to reduce the frequency of respiratory illnesses and largely decreasing absenteeism from work and medical visits ^[5]^.

Despite vaccine efficacy the coverage of many highly recommended vaccines is still inadequate and two types of barriers for vaccination have been identified: structural and attitudinal barriers ^[6]^. Structural barriers correspond to “systemic issues that may limit the ability of individual persons to access a vaccine service”, whereas attitudinal barriers refer to vaccine hesitancy or “delay in acceptance or refusal of vaccines despite the availability of vaccination services” ^[7]^.

Regarding vaccination confidence, a large-scale study in 2015 measuring worldwide variations in attitudes about vaccination, showed that the European region reported the highest mean-averaged negative responses for vaccine importance, safety, and effectiveness ^[8]^. Despite the widespread access to vaccines, France was the least confident country: 41% of French people considered vaccines to be unsafe, which represented a much higher rate than the global average (13%). Controversies related to the various vaccination campaigns in France since the 2000’s may justify the presence of anti-vaccination attitude in this country ^[9]^. For instance, a study showed that the 2009 influenza A(H1N1) crisis contributed to cause a general increase in negative attitudes towards vaccination in France ^[10]^. Nevertheless, a recent study highlighted that though vaccine confidence has been recently increasing in Europe and in France, it remains low ^[11]^.

Numerous studies showed that attitudes towards vaccination and actual vaccination behaviours are strongly associated ^[10,12–15]^. Hence, the loss of confidence in vaccine importance, safety and effectiveness of the past few years, has led to a decline in vaccination coverage against certain diseases for which vaccination is recommended by the public authorities ^[16]^, leading to vaccine preventable disease outbreaks ^[17,18]^. An illustration of this phenomenon is the measles-mumps-rubella vaccination coverage that did not reach the optimal level and thus caused a measle outbreak in France ^[19]^. In the current context of the COVID-19 pandemic, vaccination has been one of the major disease control strategies. Yet, in a study investigating vaccine acceptance amongst 33 worldwide countries, France had one of the lowest COVID-19 vaccine acceptance rates, and this could be linked to a lack of confidence in the safety of vaccines ^[20]^.

The WHO listed vaccine hesitancy as the eighth major threat for global health in 2019^[21]^ and their Strategic Advisory Group of Experts has called for better monitoring of vaccine confidence and hesitancy ^[11]^. In order to do so it is necessary to develop and validate tools. By providing insight into factors involved in vaccine hesitancy, it could enable health authorities and policy makers to address the issue with accurate messages and responses to the specific concerns of the studied populations. Existing scales are mostly aimed at parents, as a large proportion of vaccinations are given in early childhood. For example, the Parent Attitudes about Childhood Vaccines survey ^[22]^ was developed to exclusively assess parental vaccine hesitancy. Other few scales focus on specific vaccines, such as the adapted Measles Vaccine Hesitancy scale ^[23]^.

The Vaccination Attitudes Examination (VAX) scale is a brief 12-item questionnaire developed by Martin et al. ^[24]^ that assesses general attitudes towards vaccination in adults. Moreover, the VAX scale contains four subscales that enable a thorough understanding of the nature of those views: (1) mistrust of vaccine benefits, (2) worries about unforeseen future effects, (3) concerns about commercial profiteering, and (4) preference for natural immunity. A strength of the VAX scale is that its scores have been shown to be significantly associated with previous vaccination behaviours and intentions to receive future vaccines ^[24–26]^. Higher scores on all four subscales were found to be associated with higher risks of being unwilling to get a COVID-19 vaccine in 2020 ^[27]^. Therefore, the VAX scale proved to be a powerful tool to differentiate vaccinators from respondents who are more likely to refuse to get vaccinated against certain diseases.

The VAX scale has been translated and validated into several languages including Turkish ^[28]^, Spanish ^[25]^, and Romanian ^[29]^. However, there is currently no French version of the VAX scale although French is the fifth most widely spoken language in the world. Moreover, the low COVID-19 vaccine acceptance rate found in France has further highlighted the necessity to have a valid and reliable scale to evaluate general attitudes towards vaccination in a French population, especially since vaccination attitudes are strong predictors of vaccination behaviours ^[20,27]^.

Consequently, the aims of this study were (1) to translate and adapt the original English version of the VAX scale into French and (2) to test the psychometric properties of this scale in a French population-based sample of adults. In line with scale validation recommendations, we applied exploratory factor analysis followed by confirmatory factor analysis to validate the factorial structure of the French version of the VAX scale ^[30,31]^. Finally, we assessed the internal consistency, the temporal stability, and the convergent and divergent validities of the scale. We predicted that the French VAX scale would replicate the factorial structure of the original scale and would have good psychometric qualities.

## Methods

### Participants

In total, 612 participations were registered on our database, including both online and face to face participations (see the procedure section). A part of the recruitment was carried out online using ads on various social media platforms (Facebook pages, Linkedin, Instagram, and Twitter). For the rest of the recruitment, acquaintances were requested to spread the word to family, friends, students, colleagues, etc. Inclusion criteria were speaking fluently French and aged 18 years and over. There was no exclusion criterion.

### Procedure

#### The VAX scale translation process

The VAX scale have been translated and adapted into French following the backward and forward method ^[32]^. First, we contacted the developers of the VAX scale and obtained formal authorization to conduct the French translation and adaptation of the tool. Then, two independent bilingual translators translated the tool from English into the target language. A third expert identified and resolved any discrepancies between both versions into a single target translation. Finally, a bilingual translator, blinded to the initial survey items, made a back translation and a comparison between the back translation and the original version was conducted to verify that both English versions were equivalent in terms of conceptual content. Following a panel discussion, a harmonized version of the tool in the target language was proposed.

Regarding response modality, we converted the initial 6-point Likert-type scale into a 5-point Likert-type scale, ranging from “Strongly disagree” to “Strongly agree”. Indeed, scientific literature shows that odd response possibilities scale allows for the middle point to be used as a neutral point and omitting the midpoint may reduce validity when respondents lack knowledge on which to base their response ^[33,34]^.

#### Experimental design

The purpose of the study was explained to participants. Participants’ rights were also mentioned. The signature of a consent form was requested. Participants could then access the different questionnaires. These steps could be carried out either online (using Qualtrics software) or face-to-face (using paper questionnaires). In order to measure temporal validity, we invited participants to complete the VAX a second time, approximately one month (30-38 days) after the first completion.

Institution’s ethics committee approval was granted for the study (IRB Accreditation number: 202100938).

### Questionnaires

Participants were asked for basic demographic and clinical information: sex, age, education level, occupational status, whether participants have children or not, the presence of any somatic or mental chronical diseases and any medical treatment for these diseases.

#### French Version of the Vaccination Attitudes Examination (VAX) Scale

The VAX scale was originally developed by Martin et al. in 2017 ^[24]^ in English. This scale is a short tool that contains 12 statement-like items that measure general attitudes towards vaccination. For instance “I feel safe after being vaccinated”, and “Authorities promote vaccination for financial gain, not for people’s health”. In our adapted version of the scale, participants were asked to respond on a 5-point Likert-type scale ranging from “Strongly disagree” to “Strongly agree”, and items 1, 2 and 3 are reverse coded. Total scores range from 12 to 60, with higher scores reflecting stronger anti-vaccination attitudes. In addition, four subscores may be calculated by adding the following items: (a) items 1, 2 and 3 for the “Mistrust of vaccine benefit” subscale, (b) items 4, 5 and 6 for the “Worries about unforeseen future effects” subscale, (c) items 7, 8 and 9 for the “Concerns about commercial profiteering” subscale, and (d) items 10, 11 and 12 for the “Preference for natural immunity” subscale. Scores on each subscale may range from 3 to 15, whereby higher scores reflect (a) more mistrust, (b) more worries, (c) more concerns, and (d) higher preference for natural immunity, respectively.

#### Vaccination Behaviours and Intentions

As in the original validation paper ^[24]^, we collected prior vaccination behaviours and vaccination intentions to help us validate the VAX scale. Vaccination behaviours were assessed with two items asking whether respondents had received an influenza and a COVID-19 shot during the previous year. Response format was dichotomous “Yes/No”. Vaccination intentions were also assessed with two items asking whether respondents had planned to get an influenza or a COVID-19 shot in the coming year. Response format was the following “Yes/No/Maybe/Already done”.

#### Vaccine Conspiracy Beliefs Scale

The Vaccine Conspiracy Beliefs Scale (VCBS) was developed and validated in English and French by Shapiro et al. ^[35]^, to assess the presence of vaccine-specific conspiracy beliefs. It contains 7 items (e.g., “Vaccine safety data is often fabricated”) for which participants are asked to respond on a 7-point Likert scale ranging from “Strongly disagree” to “Strongly agree”. An average score is calculated, higher scores reflecting stronger conspiracy beliefs.

#### Beliefs about Medicines Questionnaire

The Beliefs about Medicines Questionnaire (BMQ) ^[36]^ assesses cognitive representations of medication. It has been validated in French in 2014 by Fall et al. ^[37]^. It consists of 18 items and participants respond on a 5-point Likert scale ranging from “Strongly agree” to “Strongly disagree”. It comprises two sections. The BMQ-Specific section contains 10 items which measure (a) the perceived necessity for a prescribed treatment (5 items e.g., “My health, at present, depends on my medicines”) and (b) potential concerns about its negative effects (5 items e.g., “My medicines disrupt my life”). By adding the reverse scores of each of the 5 items, this section allows us to calculate two scores between 5 and 25. The BMQ-General section comprises 8 items measuring (a’) responders’ beliefs about the way doctors prescribe medicines (4 items e.g., “Doctors place too much trust on medicines”) and (b’) the potential negative effects of treatment in general (4 items e.g., “Medicines do more harm than good”). By adding the reverse scores of each 4 items, this section allows us to calculate two scores between 4 and 20. Higher scores mean stronger beliefs. The two sections can be administered separately. In our study, the BMQ-Specific subscale has been administered only to participants who have previously declared that they had a chronic disease for which they needed to take a specific treatment. The BMQ-General subscale has been administered to every participant.

#### Parent Attitudes about Childhood Vaccines survey

The Parent Attitudes about Childhood Vaccines (PACV) ^[22]^ is a self-administered scale developed to assess parental vaccine hesitancy. It has been validated in French in 2021 ^[38]^. It contains 15 items with different response formats (dichotomous, “Yes/No”; 5-point Likert scale, e.g. ranging from “Strongly agree” to “Strongly disagree”, and 10-point Likert scale; e.g. ranging from “Not sure at all” to “Completely sure”). According to the method used by the initial authors ^[39]^, these three formats were then collapsed into three response categories: hesitant (receiving a score of 2), not sure/don’t know (receiving a score of 1), and not hesitant (receiving a score of zero). By adding the score of individual items, we obtained a total score ranging from 0 to 30.

#### Survey Validity check

Conducting surveys online expose researchers to more responder’s carelessness and fraud ^[40]^. Even low levels of those can lead to a decrease in the quality of collected data and to incorrect statistical conclusions. As recommended, six validated attention check items were added across the online surveys to detect careless or fraudulent responding. Four items were added to the first form and two to the second form (i.e. the form whose purpose is to test the temporal reliability of the tool, see the “procedure” section), such as “Please answer “Strongly disagree” to this question” ^[41]^. In this case, any other response than “Strongly disagree” is considered incorrect and may come from a careless or fraudulent participation. Correct answers received a score of zero and incorrect responses received a score of one. A total score out of 4 was computed for the first form and a total score out of 2 for the second form.

## Statistics and results

### Statistical analyses

Statistical analyses were carried out using IBM SPSS software version 24.0 ^[42]^. We decided to remove participants from the study based on the following criteria: not having responded to the totality of the validity-check items, having responses that did not pass the validity-check, having completed the study twice on the online survey and having abandoned the completion of the form before the end (i.e., incomplete data). The winzoring method was used to process outlier scores on our variables of interest (Field, 2018). No normal distribution was considered when absolute values for skewness and kurtosis were greater than 3 and 10, respectively ^[44]^. Means and standard deviations were computed for continuous variables and categorical variables were expressed in percentages.

An exploratory factor analysis (EFA) was conducted on the 12 items of the VAX scale. The Kaiser-Meyer-Olkin (KMO) test was used to verify the sampling adequacy. A KMO > 0.50 indicates an adequate sample to run an EFA (Field, 2018). To test intercorrelation between VAX scale items, the Bartlett’s test of sphericity was computed. If the p-value is below the 0.05 threshold, it indicates a good correlation between the variables. As our data were normally distributed, the maximum likelihood extraction method was chosen ^[45]^. The choice of the number of factors to be extracted was based on parallel analysis and scree-plot ^[46,47]^.

The parallel analysis was performed by randomly generating a data set with the same number of observations and variables as in the original data. The recommended number of factors to extract is the number of original eigenvalues that are greater than their respective 99th percentile of simulated values ^[31,45]^. A scree plot is a graph of eigenvalues against the corresponding factor numbers. The number of factors was determined by locating the point where the slope of the scree plot curve stabilises clearly below the “bend” (inflexion point). As our factors are expected to be correlated with each other, the oblimin rotation was selected. After rotation, items with loadings > 0.32 were retained and considered a statistically meaningful factor. Items with loadings < 0.32 and cross loadings > 0.32 were considered as suspect.

In order to test the factorial structure of the VAX scale obtained from the EFA, a confirmatory factor analysis (CFA) was performed using IBM SPSS Amos 26.0 ^[48]^. The model fit was assessed with the following indices: χ^2^ to degrees of freedom (χ^2^/df, p > .05), the Tucker Lewis index (TLI > 0.90), the comparative fit index (CFI > .90), and the root mean square error of approximation (RMSEA < 0.08). The model selection was made based on the Expected Cross-Validation Index (ECVI). The lower the ECVI value, the better the performance of the model ^[49,50]^.

The internal consistency of the VAX scale was assessed with McDonald’s Omega coefficient to limit the risks of overestimation or underestimation of reliability related to Cronbach’s alpha. Any values below 0.70 reflect a lack of reliability of the tool ^[51]^.

Convergent validity was examined with Pearson correlation between the VAX total score and scores of the PACV survey, the BMQ, and the VCBS. A correlation coefficient between 0.10 and 0.30 indicates a small effect size, a correlation coefficient between 0.30 and 0.50, a medium effect and a correlation coefficient above 0.50 a large effect ^[52]^. T-tests for independent samples were then conducted to compare the VAX total score between (a) participants who received an influenza vaccine during the past year and those who did not; (b) participants who received a COVID-19 vaccine during the past year and those who did not. Analyses of variance were carried out to compare the VAX total score between (c) participants who intended to get an influenza shot in the coming year, those who hesitated (i.e., “Maybe” response) and those who did not; (d) participants who intended to get a COVID-19 shot in the coming year, those who hesitated (i.e., “Maybe” response) and those who did not. Given that sample sizes were unequal for each analysis, we adjusted each sample size by randomly selecting a number of participants approximately equal to that of the smallest sample. If the analyses revealed any significant differences between groups, post hoc analyses were performed with the Bonferroni correction. Finally, the divergent validity was conducted using correlational analyses between the VAX total score and age and years of scholarship. Gender effect was explored with Student’s t-test for independent samples.

To establish the temporal stability of the VAX scale, the intraclass correlation coefficient (ICC) was computed. An ICC below 0.50 reflects poor reliability, between 0.5 and 0.75 moderate reliability, between 0.75 and 0.90 good reliability and above 0.90 excellent reliability (Cohen, 2013).

All analyses were conducted with a significance threshold of α ≤ 0.05, two-tailed.

### Results

#### Preliminary analysis

From our initial sample (n = 612), we removed 132 participants who did not respond to the totality of the validity-check items, 3 respondents who completed the study twice on the online survey and 3 respondents who abandoned the completion of the form before the end (i.e., incomplete data). Regarding the validity-check (for online participants), those who received a score > 1 (i.e. more than one wrong answer to the validity questions) were excluded from the study, representing 9% of the total sample (n = 24). In the final sample (n = 450), only 3 values were identified as outlier data, representing 0.04% of total variables. All data from questionnaires had satisfactory skewness and kurtosis values, suggesting a normal distribution.

The total sample was randomly split in two groups. The sub-sample 1 (n = 225) was used to conduct the EFA and the sub-sample 2 (n = 225) was used to conduct the CFA. Sub-samples presented no significant differences in terms of age (t_447_= 0.81, p = 0.42), sex proportion (χ^2^ = 0.29, p = 0.59) years of education (t_447_ = 0.19, p = 0.85), and experimental modality (i.e. Qualtrics vs. paper, χ^2^ = 0.04, p = 0.83).

#### Demographic and clinical characteristics

Descriptive statistics for all demographic and clinical characteristics for the total sample and the two sub-samples are presented in Table 1. Our total sample had a mean age of 32.92 years (± 14.16) and females represented 75% of our total sample group. Given that vaccine refusal and vaccine hesitancy have both been shown to be significantly associated with female sex in the literature ^[54,55]^, we decided to make an weighted adjustment based on participants’ sex. In order to do so, the proportion of females was adjusted from 75% to 50% and the proportion of males was adjusted from 25% to 50%. Weighting coefficients were 0.66 for females and 2 for males and were assigned to each variable of our dataset. All statistical analyses were then performed on the weighted data with the corresponding weighting factor.

**Table 1.**
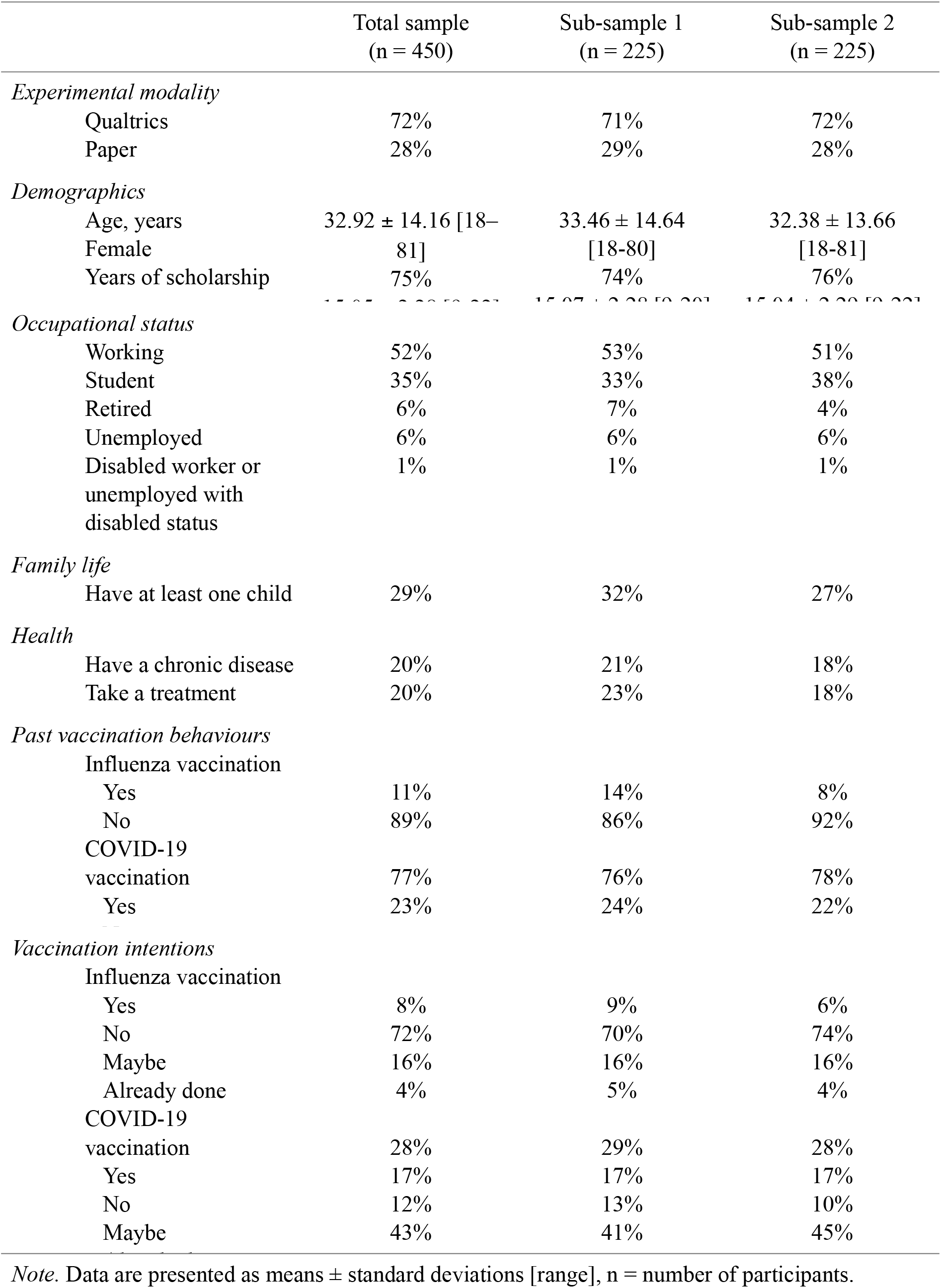
Demographic characteristics of participants.

#### Exploratory factor analysis

The EFA was conducted on sub-sample 1. Descriptive statistics of the VAX scale items and results of the EFA are documented in Table 2. Correlation matrix suggested no problem of multicollinearity. All items presented correlation coefficients > 0.30 with the other items of the scale, except for item 4 which demonstrated lower but nonetheless sufficient correlation coefficients with the other items (i.e., r > 0.16). However, we decided to keep item 4 for the next step of the analyses, as it may be clinically relevant. The overall KMO value was meritorious (0.89), and Bartlett’s test of sphericity was significant (χ^2^ = 1899, df = 66, p < .001), suggesting that the sample was appropriate to run an EFA. The parallel analysis indicated four factors with eigenvalues higher than the stimulated data (factor 1, 6.45 versus 0.61; factor 2, 0.79 versus 0.46; factor 3, 0.57 versus 0.36; factor 4, 0.46 versus 0.27). The next six factors were not supported (factor 5, 0.01 versus 0.19; factor 6, -0.00 versus 0.12; factor 7, -0.02 versus 0.06; factor 8, -0.07 versus 0.01; factor 9, -0.10 versus -0.05; factor 10, -0.12 versus -0.11) (Figure 1). The examination of the scree plot indicated a point of inflexion at five factors. The parallel analysis and the scree plot suggested both a four-factor structure, explaining 80.23% of the total variance. Based on these results, a forced four factors EFA was performed on the VAX. As found in Table 2, item 6 “*I worry about the unknown effects of vaccines in the future*” loaded both on factors 4 and 1. According to the standard recommendations, this item should be removed from the scale. However, when we removed item 6, items 4 and 5 were no longer significant. These three items referred to the dimension “Worries about unforeseen future effects”. Still, a study led during the COVID-19 pandemic showed that concerns about unknown long-term effects of vaccines were the primary reason for vaccine hesitancy (Prickett et al., 2021), reflecting the importance of retaining these items. Another study found similar results: scores of the “Mistrust of vaccine benefits” and “Worries about unforeseen future effects” VAX scale’s dimensions were found to be the most important determinants of uncertainty and unwillingness to vaccinate against COVID-19 ^[27]^. Since item 6 directly concerns the potentially harmful long-term effects of vaccines, we decided to keep it in the factor containing items 4 and 5 which also explore similar beliefs (i.e. unknown potential problems about vaccine for item 4 and potential problems for children for item 5). To conclude, we found the “Mistrust of vaccine benefits” factor, composed of items 1, 2 and 3, the “Worries about unforeseen future effects” factor, composed of items 4, 5 and 6, the “Concerns about commercial profiteering” factor, composed of items 7, 8 and 9, and the “Preference for natural immunity” factor, composed of items 10, 11 and 12. Note that, all factors are significantly correlated with each other (factor 1 and 2, r = -0.67; factor 1 and 3, r = 0.67; factor 1 and 4, r = 0.39; factor 2 and 3, r = -0.61; factor 2 and 4, r = -0.34; factor 3 and 4, r = 0.43).

**Table 2.** Mean, standard deviation and factor loadings for the Vaccination Attitudes Examination scale.

| Items | Mean ± SD | Factor 1 | Factor 2 | Factor 3 | Factor 4 |
| --- | --- | --- | --- | --- | --- |
| 1 | 2.50 ± 1.19 | 0.03 | **-0.90** | -0.03 | 0.06 |
| 2 | 1.85 ± 1.01 | 0.19 | **-0.52** | 0.18 | -0.12 |
| 3 | 2.35 ± 1.17 | -0.07 | **-1.00** | -0.02 | 0.02 |
| 4 | 3.65 ± 1.14 | -0.03 | -0.01 | -0.01 | **0.86** |
| 5 | 3.14 ± 1.18 | 0.19 | -0.05 | 0.26 | **0.46** |
| 6 | 2.70 ± 1.37 | **0.35** | -0.18 | 0.11 | **0.32** |
| 7 | 2.29 ± 1.29 | **0.79** | -0.01 | 0.04 | 0.02 |
| 8 | 2.23 ± 1.36 | **0.95** | -0.002 | -0.11 | 0.04 |
| 9 | 1.79 ± 1.22 | **0.79** | -0.04 | 0.15 | -0.08 |
| 10 | 2.88 ± 1.21 | -0.002 | -0.02 | **0.73** | -0.01 |
| 11 | 2.59 ± 1.34 | -0.03 | -0.05 | **0.86** | 0.01 |
| 12 | 2.31 ± 1.26 | 0.04 | -0.04 | **0.83** | 0.05 |
| % of variance |  | 55.41 | 9.73 | 8.09 | 7 |

*Note.* Data are presented as means ± standard deviations [range]. Factor loading at least 0.32 appear

**Figure 1.**
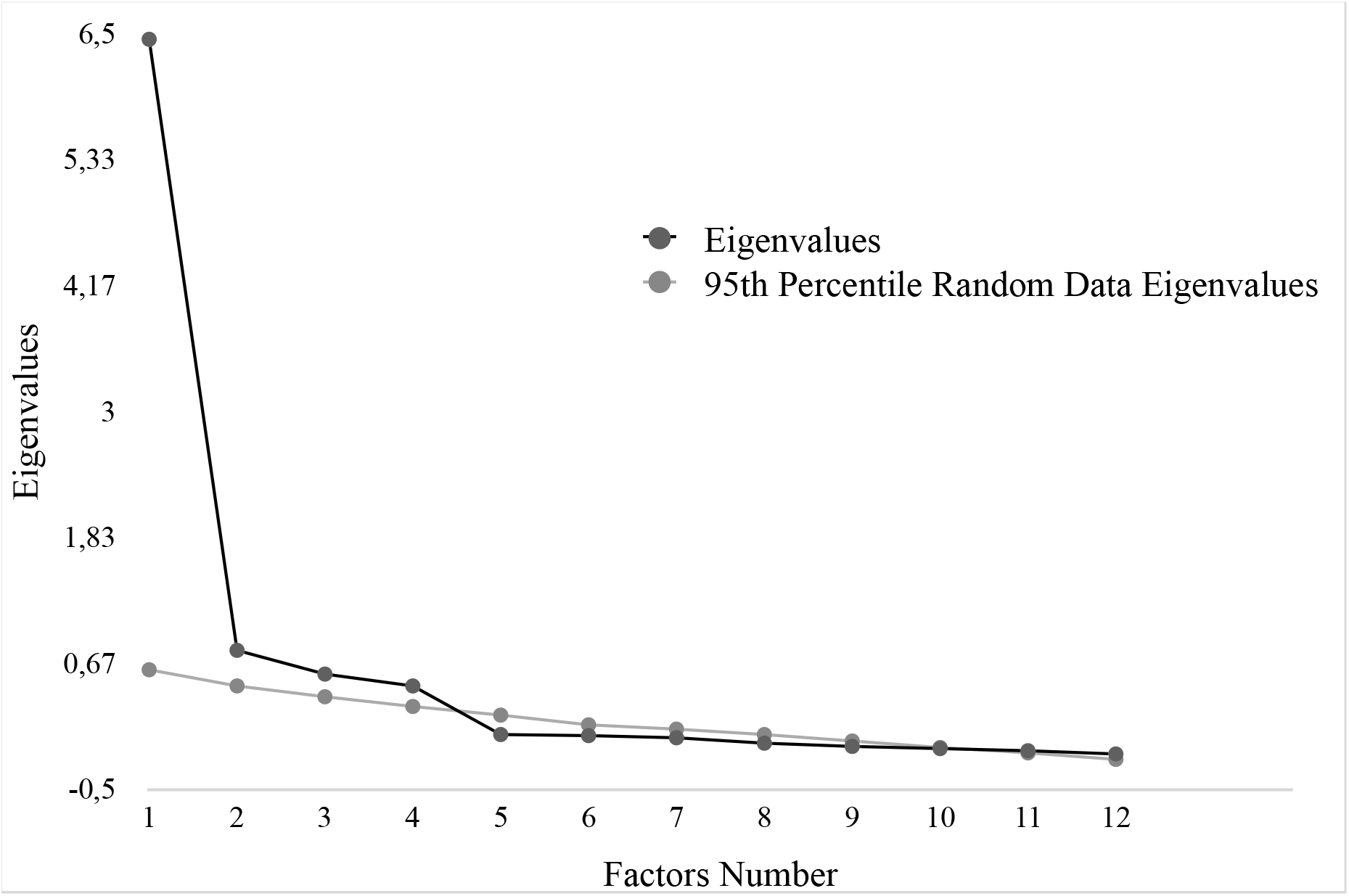
Scree Plot and parallel analysis of eigenvalues for the Vaccination Attitudes Examination factors.

#### Confirmatory factor analysis

The CFA was conducted on sub-sample 2. Two models were computed: the 4-factor model previously established with EFA, and a single-factor model (1-factor model). In the 1-factor model, all VAX scale items were an indicator of one unique latent variable. The 4-factor model showed a good fit with regards to model fit index (χ2/df = 2,65, p < .001; TLI = 0.94; CFI = 0.96; RMSEA = 0.08) while the 1-factor model showed a less satisfactory adjustment (χ2/df = 10.06, p < .001; TLI = 0.69; CFI = 0.75; RMSEA = 0.20). Moreover, comparison of the two models, based on the ECVI, showed that the 4-factor model is a better fit (4-factor model ECVI = 0.91; 1-factor model ECVI = 2.85). Figure 2 illustrates the path diagram for the retained model.

**Figure 2.**
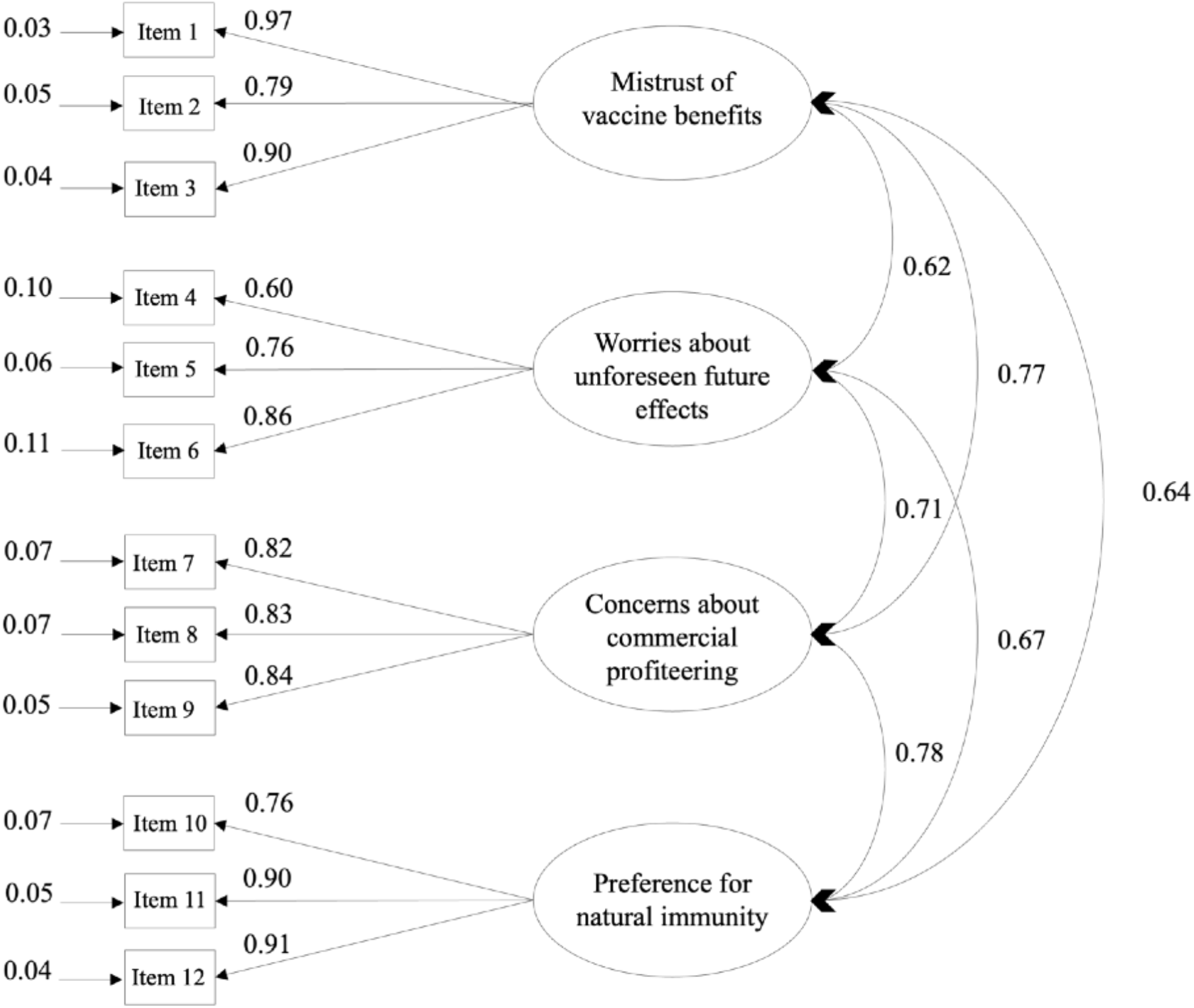
Path diagram for the for the Vaccination Attitudes Examination factors. *Note*. The four-factor model with Mistrust of vaccine benefits, Worries about unforeseen future effects, Concerns about commercial profiteering and Preference for natural immunity as latent variables. All manifest variables are represented by rectangles. Factor loadings and measurement errors are indicated by single-head arrows. The correlations between the latent variables are indicated with the double-headed arrows.

#### Internal consistency

The internal consistency was assessed using the total sample. The results revealed high internal consistency of the VAX total score (ω = 0.93) and the four factors (“Mistrust of vaccine benefit”, ω = 0.91; “Worries about unforeseen future effects”, ω = 0.79; “Concerns about commercial profiteering”, ω = 0.90; “Preference for natural immunity”, ω = 0.87).

#### Convergent validity

Means and standard deviations of questionnaires are reported in Table 3. Correlational analyses revealed that all VAX scores were strongly associated with the VCBS total score and the PACV survey total score. Moderate to strong correlations were found between all VAX scores, the BMQ-General scores and the BMQ-Concerns about its negative effects. BMQ-Perceived necessity for a prescribed treatment was moderately correlated with total VAX score and “Concerns about commercial profiteering” factor. Weak correlations were observed between “Worries about unforeseen future effects” factor, “Preference for natural immunity” factor and BMQ-Perceived necessity for a prescribed treatment. Finally, no correlation was found between “Mistrust of vaccine benefit” factor and BMQ-Perceived necessity for a prescribed treatment. Correlational analyses between questionnaires are reported in Table 4. The participants who had not received influenza and COVID-19 vaccines in the previous year had a higher VAX total score (respectively, 27.90 ± 9.15; 40.43 ± 12.38) compared to those who were vaccinated (respectively, 24.34 ± 9.09; 27.82 ± 8.64) (respectively, t_102_= 1.98, p = 0.05, d’= 0.19; t_197_= 8.31, p < .001, d’= .51). Regarding influenza vaccination intentions, a significant group effect on the VAX total score was observed (F = 14.93, p < .001, n^2^ = .23). Post hoc analyses indicated that the participants that did not plan on getting an influenza shot in the coming year had a higher VAX total score (35.57 ± 13.75) compared to those who hesitated (25.60 ± 7.22) and compared to those who did plan to get vaccinated (23.24 ± 7.16) (p < .001). There was no significant difference between participants who planned on getting an influenza shot and those who hesitated (p = 0.94). Regarding COVID-19 vaccination intentions, a significant group effect was observed on the VAX total score (F = 62.62, p < .001, n^2^ = .45). Post hoc analyses indicated that participants that did not plan on getting a COVID-19 shot in the coming year had a higher VAX total score (44.83 ± 9.33) compared to those who hesitated (31.92 ± 8.66) and did plan to get vaccinated (27.18 ± 7.68) (p < .001). Participants who hesitated to be vaccinated for the COVID-19 had a higher VAX total score compared to those who did plan to get vaccinated (p < .02).

**Table 3.** Mean and standard deviation of questionnaires.

| | Mean ± SD [min – max] |
| --- | --- |
| *Vaccination Attitudes Examination Scale*<sup>α</sup> |  |
| Total | 30.68 ± 11.01 [12 – 60] |
| Mistrust of vaccine benefits | 6.89 ± 3.22 [3 – 15] |
| Worries about unforeseen future effects | 9.64 ± 3.11 [3 – 15] |
| Concerns about commercial profiteering | 6.42 ± 3.47 [3 – 15] |
| Preference for natural immunity | 7.72 ± 3.39 [3 – 15] |
| *Vaccine Conspiracy Belief Scale*<sup>α</sup> | 2.88 ± 1.40 [1 – 7] |
| *Parent Attitudes about Childhood Vaccines Survey*<sup>β</sup> | 9.24 ± 7.93 [0 – 26] |
| *Beliefs about Medicines Questionnaire* |  |
| Specific<sup>δ</sup> |  |
| Perceived necessity for a prescribed treatment | 10.7 ± 3.93 [5 – 25] |
| Concerns about negative effects | 17.9 ± 4.19 [8 – 25] |
| General<sup>α</sup> |  |
| Beliefs about prescribe medicines | 12 ± 3.48 [4 – 20] |
| Perceived negative effects of treatment | 15.13 ± 3.23 [4 – 20] |

Note. SD = standard deviation, n = number of participants.
As the Parent Attitudes about Childhood Vaccines Survey and the Belief about Medicines Questionnaires -Specific did not cover all participants, the number of participants for these questionnaires is different from the total sample. We reported the weighted number of participants.
$\alpha$ n = 447
$\beta$ n = 129
$\delta$ n = 70

**Table 4.**
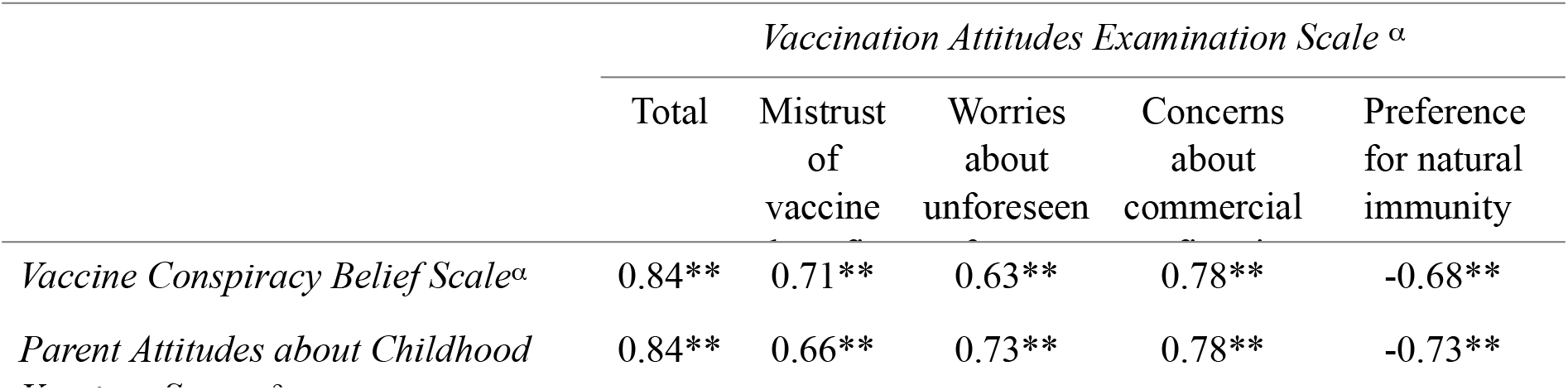
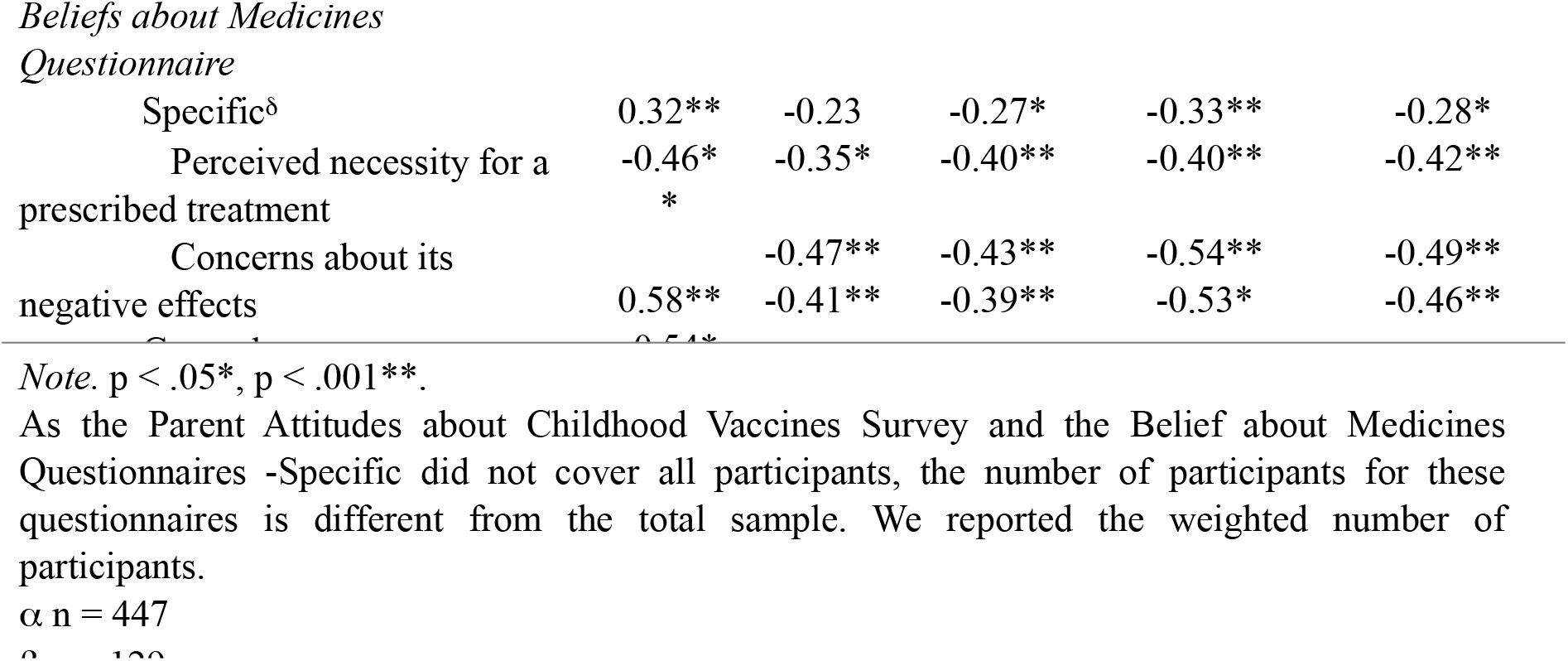
Correlational analyses between questionnaires.

##### Divergent validity

Divergent validity was assessed using the total sample. Results showed no significant correlation between age and the VAX total score (r = 0.05). A weak correlation was found between years of education and the VAX total score (r = -0.29, p < .001). Women reported a higher total score on the VAX scale than did men with a small effect size (respectively, 32.63 ± 11.34, versus men 28.65 ± 10.29, t_448_ = -3.98, p < .001, d’ = 0.18).

##### Temporal stability

A total of 168 participants completed the VAX scale twice (with a one-month interval). Descriptive statistics for both administrations are presented in Table 5. The ICC computed between the VAX total scores was significant (r =0.95 with a 95% confidence interval from 0.94 to 0.97; p < .001), reflecting excellent reliability. A paired t-test revealed that the VAX total score did not differ significantly from the first administration (28.45 ± 10.36) to the second (28.54 ± 10.23) (t_169_= -0.28, p = 0.78).

**Table 5.** Mean and standard deviation of the 1^st^ and 2^nd^ administration of the Vaccination Attitudes Examination scale.

| Items | Mean ± standard deviation | |
| --- | --- | --- |
| | 1<sup>st</sup> administration<br>Mean ± SD | 2<sup>nd</sup> administration<br>Mean ± SD |
| 1 | 2.40 ± 1.15 | 2.34 ± 1.1 |
| 2 | 1.83 ± 1.01 | 1.79 ± 0.91 |
| 3 | 2.31 ± 1.09 | 2.29 ± 1.02 |
| 4 | 3.47 ± 1.19 | 3.62 ± 1.13 |
| 5 | 3.07 ± 1.11 | 3.04 ± 1.11 |
| 6 | 2.53 ± 1.37 | 2.62 ± 1.29 |
| 7 | 2.24 ± 1.25 | 2.12 ± 1.18 |
| 8 | 2.01 ± 1.29 | 2.12 ± 1.24 |
| 9 | 1.59 ± 1.06 | 1.64 ± 1.08 |
| 10 | 2.62 ± 1.18 | 2.65 ± 1.24 |
| 11 | 2.31 ± 1.25 | 2.36 ± 1.21 |
| 12 | 2.05 ± 1.15 | 2.04 ± 1.16 |
| Total VAX score | 28.45 ± 10.36 | 28.54 ± 10.24 |
| Mistrust of vaccine benefits | 6.54 ± 2.98 | 6.41 ± 2.68 |
| Worries about unforeseen | 9.09 ± 3.10 | 9.28 ± 2.95 |
| Concerns about commercial | 5.84 ± 3.25 | 5.87 ± 3.15 |
| Preference for natural | 6.97 ± 3.17 | 6.97 ± 3.25 |
| *Note.* Data are presented as means ± standard deviations [range]. | | |

## Discussion and limitations

The goals of the present study were (1) to translate and adapt the original English version of the Vaccination Attitude Examination (VAX) scale into French and (2) to test the psychometric properties of this scale in a French population-based sample of adults.

As hypothesized, the exploratory factor analysis showed a 4-factor solution with 12 items that explained 80.23% of the variance. The four factors are: “Mistrust of vaccine benefits” (items 1, 2, and 3), “Worries about unforeseen future effects” (items 4, 5, 6), “Concerns about commercial profiteering” (items 7, 8 and 9), and “Preference for natural immunity” (items 10, 11 and 12). Moreover, the French adaptation of the VAX scale demonstrated high internal consistency reliability and a strong temporal stability.

Further statistical analyses provided more arguments to establish validity, in particular convergent validity. The literature shows that endorsing vaccine conspiracy beliefs predicts general attitudes towards vaccine ^[56,57]^. This relationship is verified in our study since our correlational analyses showed a positive correlation between total scores of the VAX scale and those of the Vaccine Conspiracy Beliefs Scale. We also found positive correlations between total scores of the VAX scale and all those of the Beliefs about Medicine Questionnaire (BMQ) subscales. These findings are in line with previous conclusions of a replication study assessing validity of the VAX scale and the Spanish validation of the scale ^[25,26]^. Indeed, the perceived necessity for a treatment and eventual concerns about the adverse consequences of it has been identified as playing a role in patients’ attitudes and decisions about treatment ^[58,59]^. We also tested the relationship between the total scores of the VAX scale and those of the Parent Attitudes about Childhood Vaccines (PACV) survey total score which represents another measure of general attitudes towards vaccination. As expected, we found a positive correlation between the two scores. This correlation replicates results found in the original validation paper ^[24]^.

Attitudes towards vaccination and actual vaccination behaviours have been found to be strongly associated in numerous studies ^[10,12–15]^. This is reflected in our results since we demonstrated that total score of the VAX scale successfully differentiated both COVID-19 and influenza vaccinators from non-vaccinators. In other words, participants who did not receive these vaccines in the previous year manifested stronger anti-vaccination attitudes than those who did. A similar pattern was found for vaccination intentions: participants who did not plan on getting vaccinated against influenza in the coming year manifested stronger anti-vaccination attitude than those who did and those who hesitated. The relationship between attitudes towards vaccination and vaccination intentions is even stronger regarding the COVID-19 vaccine since the VAX scale is able to differentiate all three groups of respondents: those who planned on getting vaccinated, those who did not, and those who hesitated. These results highlight that the VAX scale is a powerful tool to differentiate vaccinators from respondents who are more likely to refuse to get vaccinated against certain diseases. Moreover, our results replicate the findings of the other VAX scale validation papers^[24–26]^.

In terms of divergent validity, total scores of the VAX scale are unrelated to age of the participants. Regarding sex effect, it appears that females reported stronger antivaccination attitudes than males. This is in line with recent findings in the scientific literature ^[20,54,55]^. Even though the VAX scale scores was unrelated to level of schooling in the original validation paper ^[24]^, a small correlation was also found between educational level and total scores of the VAX scale. However, the authors mentioned that their results may be related to the fact that the range on education was restricted in their sample. Our result is not surprising since such findings have already been found in other studies: more vaccine refusal and vaccine hesitancy seem to be associated with lower educational levels ^[54,55]^. Moreover, in the replication validation study conducted by Wood et al in 2019 ^[26]^, the relation between VAX score and education level is also found: increased education predicted lower VAX scores.

Nevertheless, the results of this study should be considered carefully. The first limitation is that our sample is mostly composed of women. Although conducting surveys online appears to be a simple mean of data collection, women tend to be more likely to select themselves into research projects ^[60]^. However, we minimised this bias by making a weighted adjustment of our data based on participants’ sex. Secondly, the current pandemic context introduced another bias: although our scale is supposed to assess general attitudes towards vaccination, vaccination is currently a worldwide hot topic. Thus, there is a risk that participants responded to our survey in reference to COVID-19 vaccine only, even though the emphasis was placed on this distinction during the face-to-face interviews; but this was not possible for online participants. Another limit of this study is that we did not test whether this factor structure held for clinical populations.

To conclude, the French version of the VAX scale is a simple tool that demonstrated strong results in the various statistical analyses carried out to validate the tool. The validation of the French VAX scale enables researchers to identify groups of particular concern with respect to vaccine hesitancy and to determine specifically which aspect of vaccination is especially problematic among these groups. The French population is particularly affected by anti-vaccination attitudes and vaccine refusal and this tool allows to study the dynamics involved in this health issue. A further understanding of attitudes towards vaccination will enable healthcare institutions to directly address populations’ specific concerns about vaccination and therefore, improve vaccine acceptance rates as a beneficial consequence.

## Data Availability

All data produced in the present study are available upon reasonable request to the authors

## Acknowledgments

The authors are grateful to all participants for the time they took to participate to this study.

